# Postprandial changes in gastrointestinal function and transit in cystic fibrosis assessed by magnetic resonance imaging

**DOI:** 10.1101/2020.02.19.20022020

**Authors:** C. Ng, N. S. Dellschaft, C. L. Hoad, L. Marciani, L. Ban, A. P. Prayle, H. L. Barr, A. Jaudszus, J. G. Mainz, R. Spiller, P. Gowland, G. Major, A.R. Smyth

## Abstract

**Background and Aims:** Cystic fibrosis (CF) is a multi-system genetic disorder affecting >72,000 people worldwide. Most people with CF experience gastrointestinal symptoms and some will develop complications such as distal intestinal obstruction syndrome. However the mechanisms of symptoms and complications are not understood. We evaluated gut function and transit of CF using magnetic resonance imaging (MRI). Our hypotheses were: oro-caecal transit time (OCTT) is longer in CF, with lower small bowel water content (SBWC).

**Methods:** Twelve people with CF at a tertiary centre and 12 age and sex-matched controls underwent serial MRIs over 1 day, with meals at set times. The primary endpoint was OCTT, assessed by the appearance of a food bolus in the caecum. Other measures included SBWC, colonic volume, gastric half-emptying time and gastrointestinal symptoms.

**Results:** OCTT was longer in CF (controls 210 minutes [173, 315] vs. CF 330 minutes [270, >360], p=0.04). There was no difference in gastric half-emptying times (controls 80 minutes [66, 88] vs. CF 97 [71, 128], p=0.3). Corrected SBWC was higher in CF (controls 34 L.min/m^2^ [28, 41] vs. CF 63 L.min/m^2^ [36, 80], p=0.021), with minimal second post-prandial decrease suggesting impaired ileal emptying. Corrected colonic volumes were higher in CF (controls 123 L.min/m^2^ [89, 146] vs. CF 186 L.min/m^2^ [166, 209], p=0.012). There were no differences in gastrointestinal symptoms.

**Conclusions:** Significant differences in gut function and transit exist between CF and controls. Our methodology provides a platform for studying gastrointestinal function in CF and has identified new potential mechanisms of dysfunction.

ClinicalTrials.gov NCT03566550

## Background and Aims

Cystic fibrosis (CF) is an autosomal recessive disorder, affecting over 72,000 people worldwide.^1,2^ The mutation with the highest prevalence is p.Phe508del and between 85-90% of people with CF have at least one copy of this gene mutation.^3,4^ The genetic defect leads to a dysfunctional CF transmembrane conductance regulator (CFTR) protein which disrupts the passage of chloride and bicarbonate ions, causing increased viscosity of epithelial mucus. CFTR is expressed on many epithelia, including the respiratory and gastrointestinal systems, which are most affected in CF.^1,5^ Nearly 20% of new CF diagnoses are made following meconium ileus in the newborn, where obstruction at the terminal ileum can lead to volvulus, ischaemic necrosis and perforation requiring surgery.^6^

Respiratory problems have historically been the main cause of premature death in CF but improvements in treatment, and thus life expectancy, have focused attention on factors affecting quality of life and extra-pulmonary involvement. In 2018, a James Lind Alliance Priority Setting Partnership involving patients, their families and clinicians identified the relief of gastrointestinal symptoms such as stomach pain, bloating and nausea as one of the top priorities for research in CF.^7^

Gastrointestinal symptoms affect the majority of people with CF.^8^ In any one year, nearly 50% report constipation and up to 20% report gastrointestinal complications, such as gastro-oesophageal reflux, distal intestinal obstruction syndrome and rectal prolapse.^3,9^ Recent efforts to describe comprehensively the burden of gastrointestinal symptoms in CF, such as the recent GALAXY study^10^ in North America, have used questionnaires derived for use in functional bowel disorders such as the Patient Assessment of Constipation-Symptoms (PAC-SYM)^11^ and Patient Assessment of Gastrointestinal Disorders Symptom Severity Index (PAGI-SYM) for upper gastrointestinal symptoms.^12^ In the United Kingdom, use of the Gastrointestinal Symptoms Rating Scale (GSRS) has identified a high symptom burden in a large proportion of patients.^13^ CF-specific questionnaires such as the CFAbd-Score have also been derived and validated^14,15^. Such work can describe the prevalence and impact of symptoms but does not provide an understanding of the mechanisms by which CTFR dysfunction impacts upon gastrointestinal function.

The mechanisms of gastrointestinal dysfunction in CF are not fully understood as there has been limited research into its pathophysiology. It has been proposed that CFTR-related impaired composition of secretions, dysmotility, dysbiosis and intestinal inflammation contribute to CF gut dysfunction.^16-19^ However, there have been no studies linking symptoms with physiological disturbance of gut function in CF. Invasive physiological measurements in the gut (such as endoscopy) are problematic in CF, as patients already have a high procedural burden. Furthermore, radiological imaging will increase further the lifetime radiation exposure of patients already subject to regular chest radiographs and CT scans. Capsule technologies for physiological assessment are not approved for use in children.

Magnetic Resonance Imaging (MRI) has been used to study gut function non-invasively for over 20 years. The use of validated MRI measures of gastric emptying,^20^ small bowel water content (SBWC),^21^ oro-caecal transit time (OCTT),^22^ and colonic volumes^23^ have been applied to gut diseases such as irritable bowel disease, inflammatory bowel disease and constipation. These measures could increase our understanding of the relative importance of factors such as the viscosity of gut content and dysmotility, in the lower intestinal pathophysiology of CF.

### Aims

The aim of the study was to evaluate the use of MRI to assess gut function and transit in people with CF. Our principal hypothesis was that OCTT would be slower in people with CF than healthy controls. Our secondary hypotheses were that:

- People with CF would have less small bowel water than controls due to inhibition of intestinal chloride secretion through CFTR dysfunction
- People with CF would have larger colonic volumes because of the prevalence of constipation in CF.
- Symptoms in CF would correlate with underlying physiological abnormalities

## Materials and Methods

### Study design

We conducted a prospective observational study, using MRI to compare the postprandial changes in gut function and transit in people with and without CF. All study procedures took place at the Sir Peter Mansfield Imaging Centre, Nottingham, UK.

### Study population

The diagnosis of CF was established in line with the CF Foundation consensus recommendations.^24^ All participants with CF were homozygous for the p.Phe508del mutation and aged between 12 and 40 years. Participants with CF were recruited from the paediatric and adult CF service at Nottingham University Hospitals NHS Trust, a tertiary centre. Participants with CF were approached in order of outpatient clinic or ward attendance to reduce recruitment bias. Participants with CF were not required to have a history of gastrointestinal symptoms or complications. Controls, who had no clinical evidence or suspicion of CF, were recruited by open advertisement in the Nottingham area and were matched to cases by age and gender.

We applied the following exclusion criteria for all participants:

- Measurement of forced expiratory volume in 1 second <40% predicted using the Global Lung Initiative criteria (for patients with CF).
- Contra-indication to MRI scanning.
- Unable to stop medications prescribed to alter bowel habit.
- Previous resection of any part of the gastro-intestinal tract apart from appendicectomy, cholecystectomy. Patients could be included if they had undergone gut resection for neonatal meconium ileus or for distal intestinal obstruction syndrome provided ≤20cm of bowel was resected.
- Intestinal stoma.
- Diagnosis of inflammatory bowel disease or coeliac disease confirmed by biopsy.
- Gastrointestinal malignancy.
- Inability to comply with dietary restrictions or tolerate meals required for the study.

### Procedures

Participants spent one day at the Sir Peter Mansfield Imaging Centre, Nottingham, UK. They fasted from 20:00h the previous evening, other than a glass of water for essential medicines. On the day before scanning, participants avoided strenuous exercise and certain food and drink that could affect bowel habit including: caffeine, alcohol, dietary or sports supplements, beans, peas, lentils and sweetcorn.

Participants stopped taking any laxatives or anti-diarrhoeals on the day of scanning. Participants completed two symptom questionnaires, the PAC-SYM^11^ and CFAbd-Score,^14^ regarding symptoms during the preceding 2 weeks. They then underwent a fasting baseline set of MRI scans, followed by their first meal. Post-prandial MRI scans were performed immediately after the meal, then at 30 minutes intervals for 180 minutes with further scans at 240, 300 and 360 minutes, giving 11 scans in total (Figure 1). After each scan, patients scored their degree of abdominal pain, bloating and flatulence on a Likert scale ranging between 0 (not at all) and 3 (severe/disabling), as used in the GSRS.^13^ A second meal was eaten after the scan at 240 minutes.

**Figure 1.**
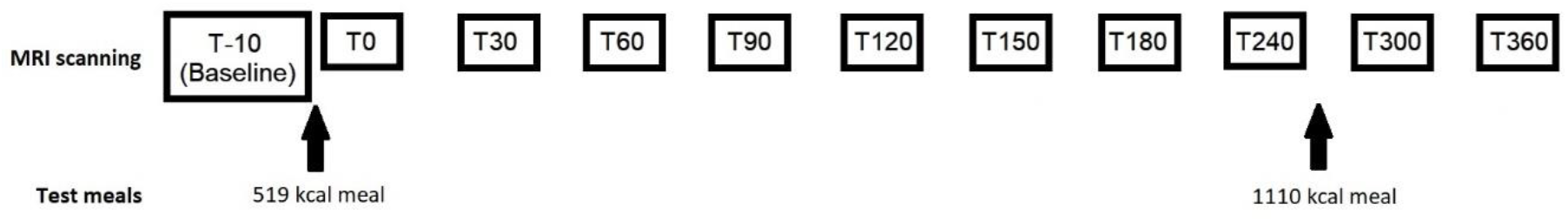
MRI study day schedule. Test meals are same standardised meals given to each participant. “T” is the time related to the when the first test meal was eaten. For example, T0 is the MRI scan taken at 0 minutes after the first meal was eaten.

No food or drink was permitted during the scan day other than the standardised meals provided. The first meal consisted of 300 g creamed rice pudding uniformly mixed with 25 g seedless raspberry jam and 30 g double cream, and a drink of 100 mL orange juice with 240 mL water (all Sainsbury’s^®^, UK). Total energy content was 519 kcal (fat 19 g and carbohydrate 77 g). The second meal consisted of 400 g macaroni cheese (Sainsbury’s^®^, UK), 100 g strawberry cheesecake (Rhokett^®^, UK) and 240 mL water. Total energy content was 1110 kcal (fat 54 g and carbohydrate content 116 g). Participants with CF took their usual dose of pancreatic enzyme replacement therapy with meals. A comfortable waiting area was available for use between scans.

### MRI scanning protocol

MRI scanning was performed on a 3T Philips Ingenia scanner (Philips Healthcare, Best, The Netherlands). Participants were positioned supine with the DS anterior coil over the abdomen in order to image the stomach, small and large bowel. A plastic guard held the coil away from the participant’s chest to allow for optimal breathing and a respiratory belt was used to monitor breathing patterns. An initial survey scan was acquired to determine location of the abdominal organs, followed by the following sequences:

1. To identify OCTT and measure colon volumes: a dual fast field gradient echo sequence with echo times 1.15 ms and 2.30 ms, repetition time 110 ms and 60° flip angle. 22 coronal slices (7 mm thick with 1 mm gap) were acquired with voxel size 1.6 × 1.6 × 7 mm.
2. To aid identification of the head of the meal at the caecum for OCTT analysis: a high resolution balanced turbo field echo sequence with echo time 1.2 ms, repetition time 2.4 ms and 42° flip angle. 8 sagittal slices (7 mm thick with 0.6 mm gap) were acquired with voxel size 1.5 × 1.5 × 7 mm.
3. To assess gastric emptying time: a half Fourier turbo spin echo sequence with echo time 60 ms, repetition time 508 ms and 90° flip angle. 20 transverse slices (10 mm thick with no gap) were acquired with voxel size 1.59 × 1.99 × 10 mm.
4. To assess small bowel water content (SBWC): a strongly T2-weighted turbo spin echo sequence (rapid acquisition with relaxation enhancement) with echo time 400 ms, repetition time 1263 ms and 90° flip angle. 24 coronal slices (7 mm thick with no gap) were acquired with voxel size 1.40 × 1.76 × 7 mm.

All sequences were run as series of breath holds, with the maximum breath hold time of 10 s, and acquired at 11 different time points throughout the day (Figure 1). Participants spent up to 15 minutes inside the magnet for each time point and spent the rest of the study day upright in an adjacent room.

### Data analysis and masking

MRI analyses were carried out using Medical Image Processing, Analysis and Visualisation (MIPAV, NIH, Bethesda)^25^ and in-house software written in IDL^®^ 6.4 (Research Systems Inc. Boulder, Colorado, USA). MRI data were relabelled by an independent researcher to ensure the researchers performing the MRI analyses were unaware of which images related to specific participants.

### Outcome measures

The primary outcome measure was OCTT. This was determined by the time of the first scan at which the head of the meal was detectable in the caecum on dual echo sequences,^22,26^ as demonstrated in Figure 2. OCTTs were checked by two researchers, with review by a third researcher to resolve any disagreements.

**Figure 2.**
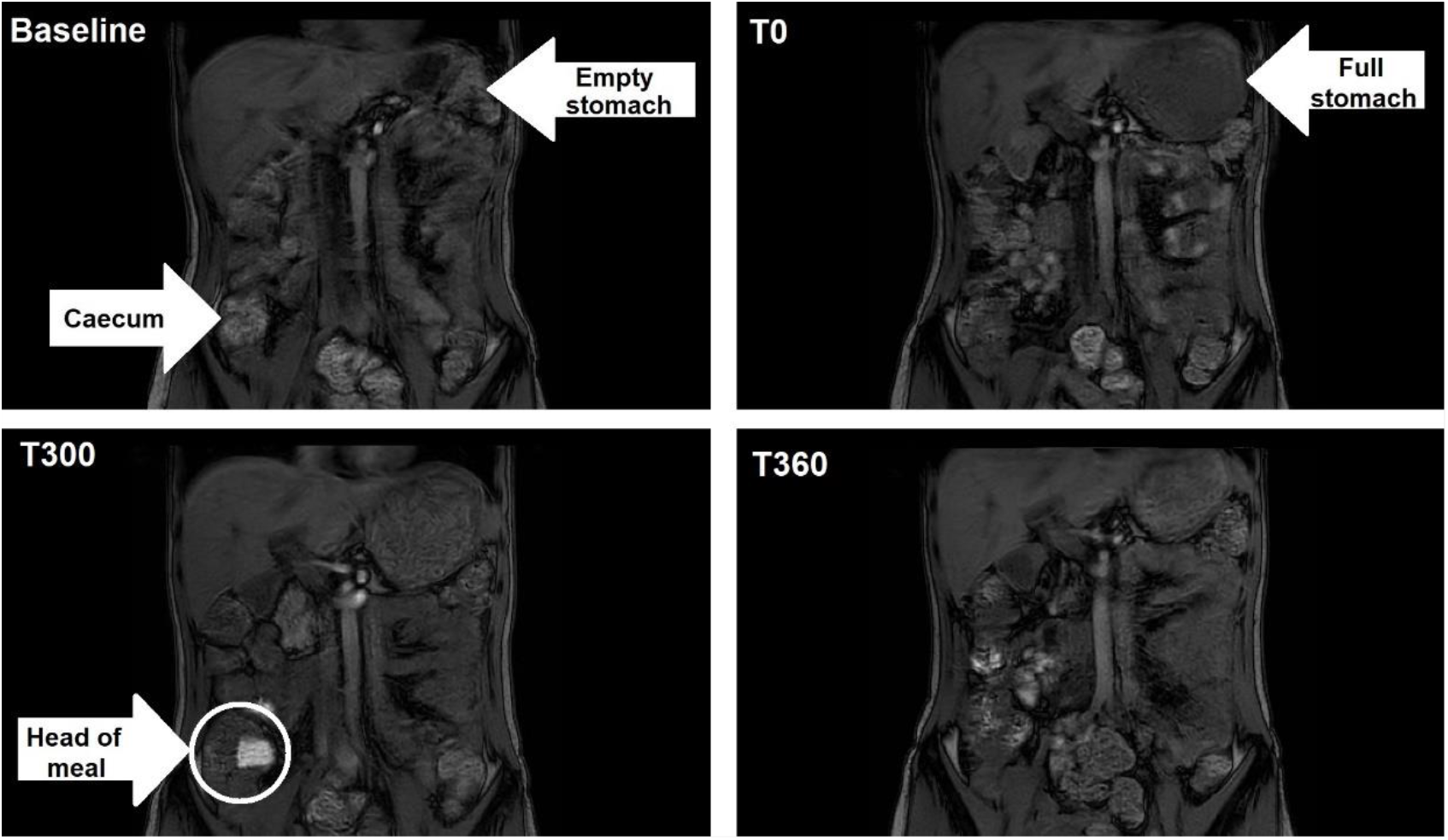
MRI images to show the head of the meal arriving in the caecum in one participant, identified by high intensity material at T300. Baseline: fasting scan. T0: scan taken immediately after first meal. T300: scan taken at 300 minutes following the first meal. T360: scan taken at 360 minutes following first meal.

Secondary outcomes included: colonic volume corrected for body surface area (area under the curve, AUC); SBWC corrected for body surface area (AUC); gastric volumes for gastric half-emptying time; and gastrointestinal symptom scores. Body surface area was calculated using Mosteller’s formula (√ [(height in cm × weight in kg)/3600]).^27^

Gastric^20^ and colonic volumes^23^ were determined as described previously by manually drawing regions of interest around the organs using imaging software (MIPAV, NIH). Gastric volumes were measured at each scanning time point until T240 (Figure 1). The time taken for the stomach to half empty calculated (gastric half-emptying time) from sequential volumes, until the volume fell below 100 mL, at which point no further volumes were included.^28^

Colonic volumes were determined for the ascending, transverse, descending and recto-sigmoid segments.^23^ When segments were fully collapsed and therefore not visualised on MIPAV, a volume of 0 mL was assigned. A random ten percent of scans were reviewed by a second researcher. The limit of inter-observer variability of no more than 5% in keeping with previous data.^29^ For each participant, the colonic volume AUC over the scan time period was calculated and divided by the body surface area to form the corrected colonic volume.

SBWC was analysed using validated in-house software as previously described.^21^ The intensity threshold for free water was set using each participant’s spinal fluid, and only small bowel contents having an intensity equal to or greater than this value was included as small bowel free water volume.^21,26^ For each participant, the SBWC AUC over the scan time period was calculated and divided by the body surface area to form the corrected SBWC.

Gastrointestinal symptoms over the 2 weeks preceding the scan day were assessed using the CFAbd-Score^14^ and PAC-SYM^11^ questionnaires as described above. The CFAbd-Score comprises 28 items grouped into 5 domains (pain, gastro-oesophageal reflux, disorders of bowel movement, eating and appetite, impairment of quality of life) which are weighted to provide a final score.^14^ The PAC-SYM final score is determined by the total score divided by the number of items answered.^11^ For both questionnaires, a lower score correlates with lower symptom burden.

Symptom scores at each MRI time point were calculated from the sum of Likert scale component scores for abdominal pain, bloating and flatulence.^13^

### Statistical analysis

This was a pilot study and so there was little prior data on which to base a power calculation.^30^ Non-parametric analyses (Wilcoxon signed-rank test) were planned for corrected colonic volumes, corrected SBWC, gastric half-emptying times, and gastrointestinal symptom scores because of the matched study design and small sample size. For OCTT, Kaplan-Meier curve and a Log rank test were used. Statistical analysis was carried out using RStudio, Inc (version 1.1.463, Boston). A p-value of less than 0.05 was considered significant.

### Ethical Considerations

The study was approved by the UK National Research Ethics Committee (18/WM/0242) and all participants gave written informed consent. For participants aged 15 years and under, written assent and parental consent were given. The study was registered on a publicly accessible trials database prior to commencement (ClinicalTrials.gov NCT03566550, Supplementary file). All authors had access to the study data and reviewed and approved the final manuscript.

## Results

### Study Progress

All procedures took place between August 2018 and February 2019. There were 51 eligible participants with CF, of whom 13 declined to participate and 2 withdrew consent. The remaining 36 eligible participants with CF were approached in order of their hospital attendance and first 12 to provide written consent were enrolled into the study. Twelve age and gender-matched controls were also enrolled.

All 12 people with CF tolerated the standardised test meals and MRI scanning without any adverse events. Twelve controls completed the protocol. One further control participant was withdrawn and replaced as they were unable to complete the first standardised meal and the remaining 10 MRI scans.

Although participants were age and gender matched, controls tended to be taller and heavier than people with CF, reflecting the nutritional challenges and impaired growth in people with CF (Table 1). Therefore, we corrected SBWC and colonic volumes for body surface area to adjust for these factors.

**Table 1.**
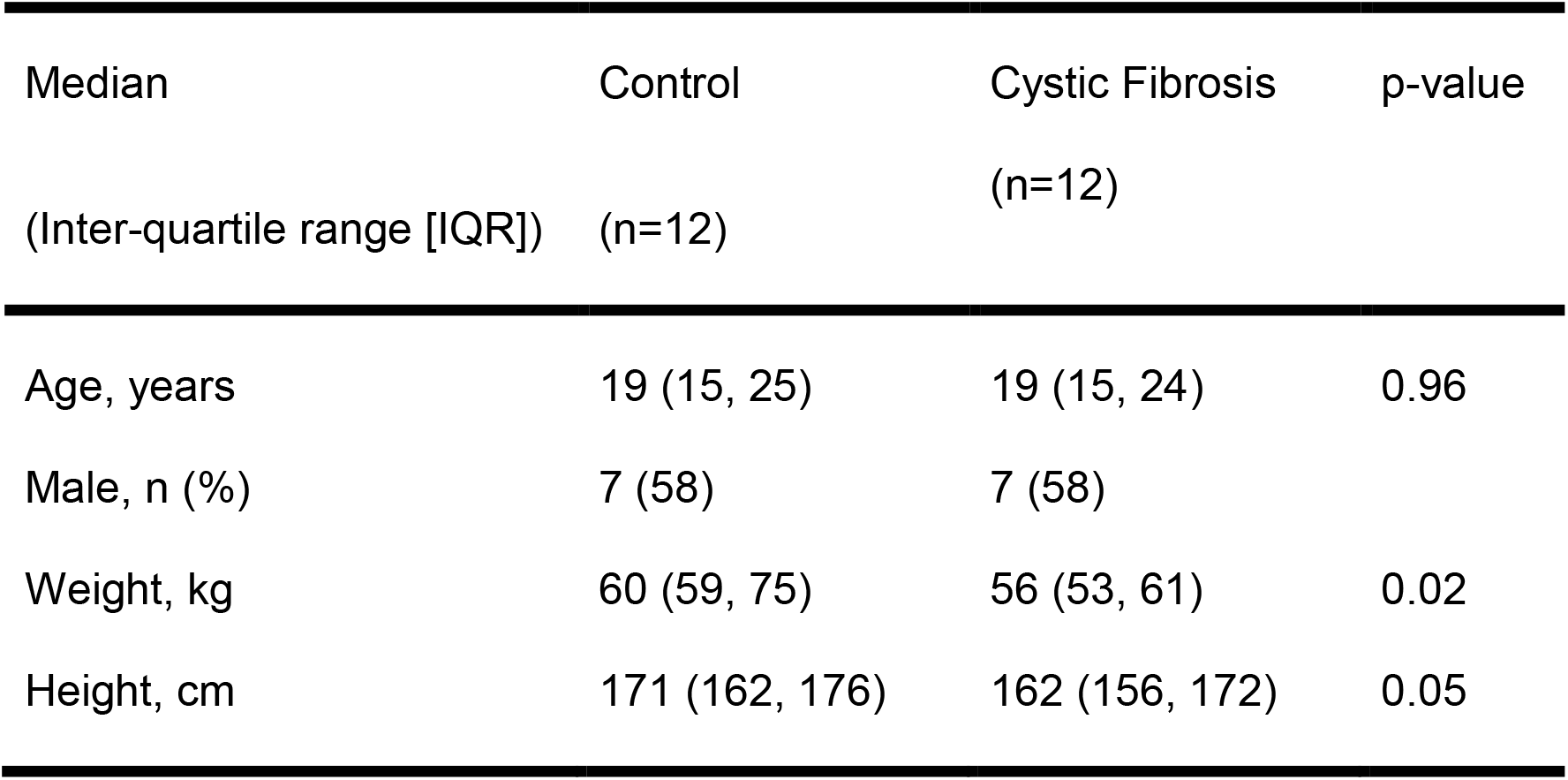
Baseline characteristics for control and CF groups at the time of MRI scanning.

### Primary Outcome

#### Oro-Caecal Transit Time (OCTT)

Transit times were significantly longer in CF cases than controls, with a difference in median OCTT of 2 hours (controls 210 minutes [IQR 173, 315] vs. CF 330 minutes [IQR 270, >360], p = 0.04). An upper limit of detection was imposed by the timing of the last scan, 360 minutes post-prandial. By this point, the meal had still not reached the caecum in 4 people with CF, suggesting an OCTT greater than 360 minutes. This was confirmed by the high intensity of the meal still seen within the small bowel at 360 minutes. All controls had an OCTT of 360 minutes or less.

### Secondary outcomes

#### Gastric half-emptying time

There was no difference in the median gastric half-emptying times between the groups (controls 80 minutes [IQR 66, 88] vs. CF 97 minutes [IQR 71, 128], p = 0.3, Wilcoxon).

#### Corrected Small Bowel Water Content (SBWC)

Corrected values for SBWC were higher in people with CF than controls. This was contrary to our *a priori* hypothesis. While no difference in baseline SBWC was found (controls median 102 mL/m^2^ [IQR 65, 120] vs. CF median 135 mL/m^2^ [IQR 90, 145], p = 0.73, Wilcoxon), the median AUC for SBWC was higher in CF (controls 34 L.min/m^2^ [IQR 28, 41] vs. CF 63 L.min/m^2^ [IQR 36, 80], p = 0.021, Wilcoxon, Figure 3).

**Figure 3.**
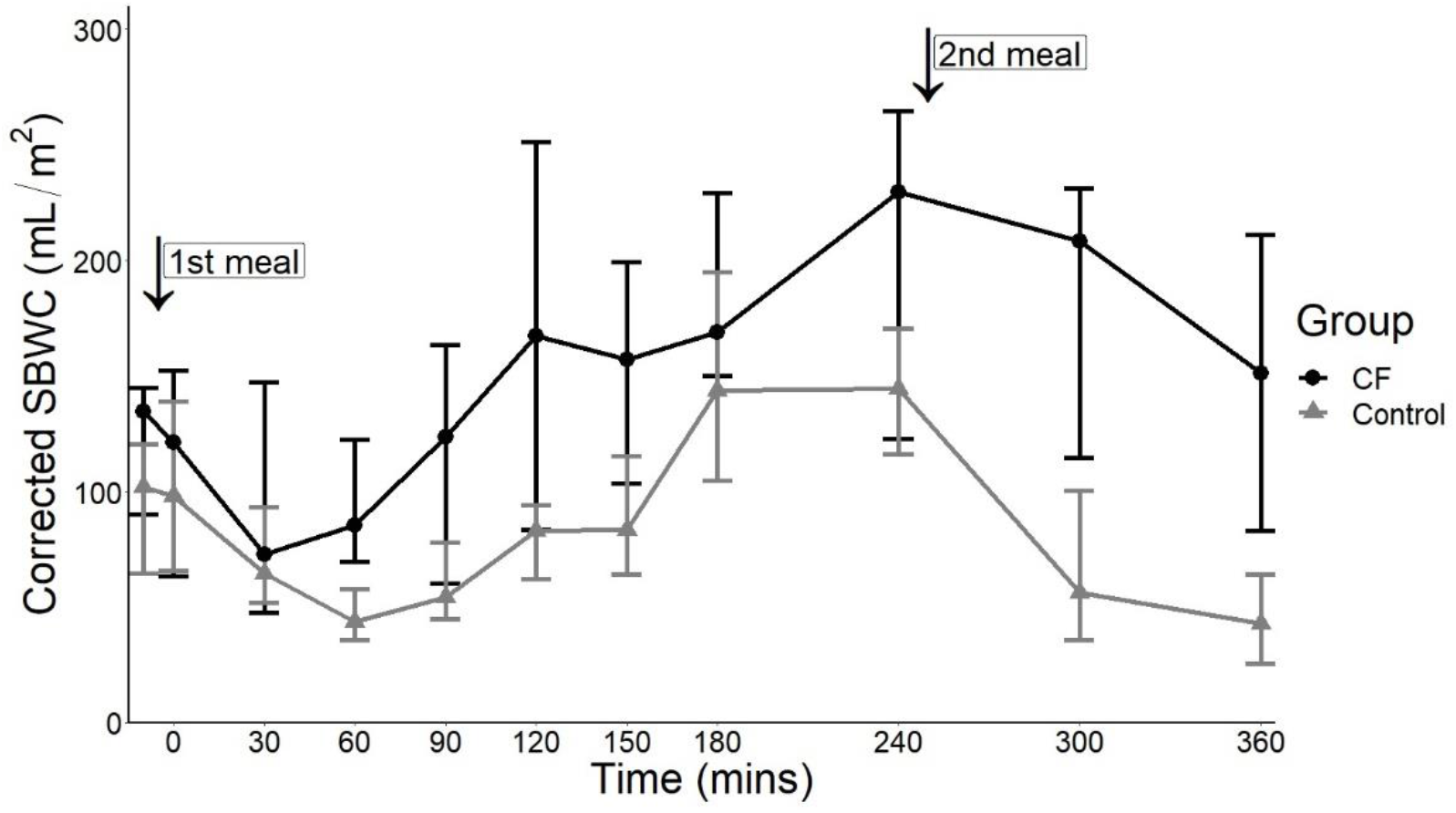
Corrected SBWC at each scanning time point for control and CF group. Error bars show median and IQR. Between 240 and 300 minutes, the fall in corrected SBWC is reduced in the CF group compared to the control group.

Further observation of the SBWC showed that the CF group did not show the large fall in SBWC after the second meal that is usually observed in a healthy group (T240 to T300 scans). Exploratory analysis showed there was only a minimal decrease in median SBWC in the CF group (CF 13 mL/m^2^ [IQR −13, 57] vs. controls 101 mL/m^2^ [IQR 67, 106], p = 0.002, Wilcoxon).

#### Corrected Colonic Volumes

Corrected baseline total colon volumes, composed of the sum of the ascending, transverse, descending and recto-sigmoid segments, were not statistically significantly different between the median of the two groups (controls 387 mL/m^2^ [IQR 352, 482] vs. CF 542 mL/m^2^ [IQR 454, 711], p = 0.092, Wilcoxon). The baseline recto-sigmoid colon was not visualised in one control. However, there was a significant difference of 63 L.min/m^2^ in corrected total colon volume AUC between the medians of the control and CF group (controls 123 L.min/m^2^ [IQR 89, 146] vs. CF 186 L.min/m^2^ [IQR 166, 209], p = 0.012, Wilcoxon, Table 2, Figure 4). The transverse colon at T360 for one control and the recto-sigmoid at 6 time-points for another control were not visualised.

**Table 2.** Corrected colonic volumes for control and CF group

| Corrected colon volume (AUC),<br>L.min/m<sup>2</sup> (Median, IQR) | Control | Cystic Fibrosis | p-values |
| --- | --- | --- | --- |
| Ascending colon | 35 (29 - 46) | 57 (42 – 71) | 0.001 |
| Transverse colon | 40 (23 – 49) | 54 (41 – 67) | 0.03 |
| Descending colon | 22 (18 – 31) | 38 (31 – 46) | 0.05 |
| Recto-sigmoid colon | 20 (18 – 27) | 45 (33 – 50) | 0.04 |

**Figure 4.**
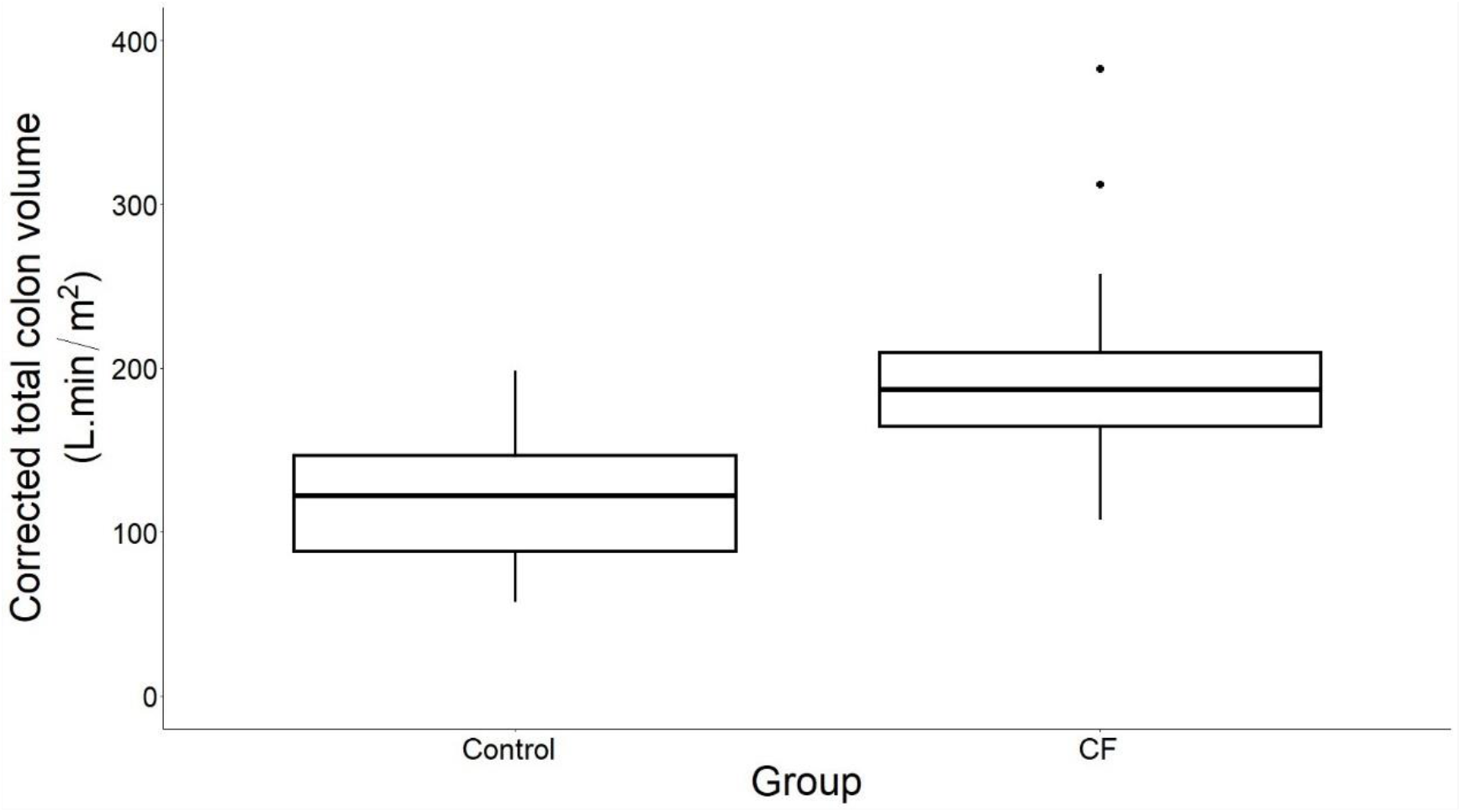
Box and whisker plot indicating the median, interquartile range and outliers of the corrected total colon volumes (AUC) for the control and CF group.

### Symptoms

#### CFAbd-Score

Total CFAbd-Score tended to be higher in CF than the control group but not statistically different (controls median 7 [IQR 3, 14] vs. CF 16 [IQR 5, 25], p=0.13, Wilcoxon) and there was no correlation with our primary and secondary MRI outcomes. The domains: pain, disorders of bowel movement and impairment of quality of life (QoL) scored numerically higher in the CF group but not statistically different compared to the control group (Figure 5).

**Figure 5.**
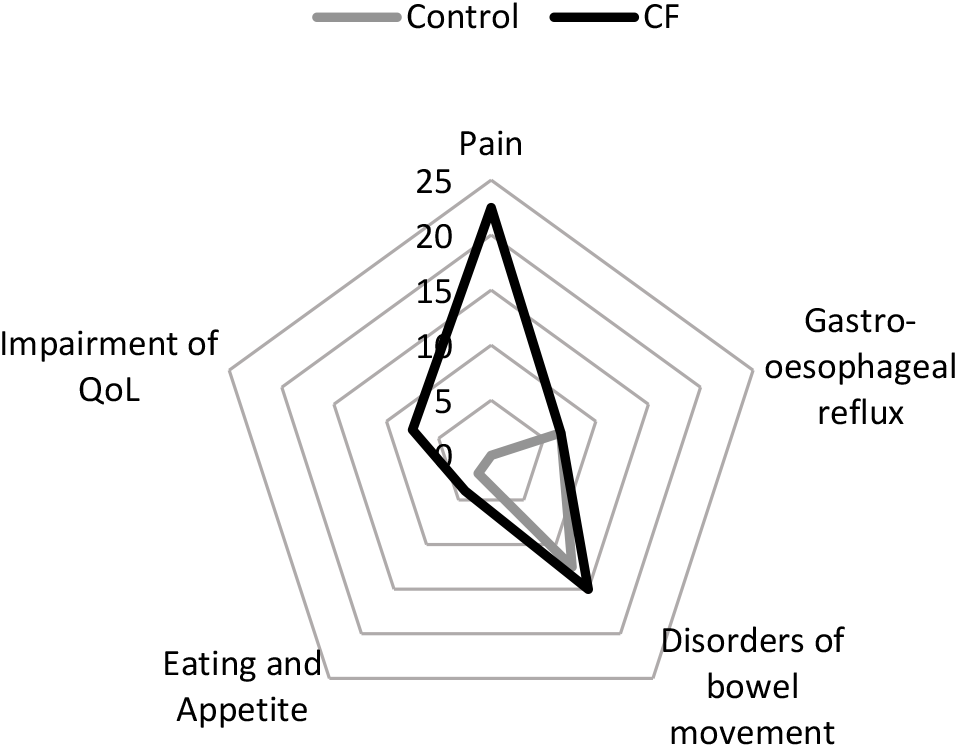
Median domain scores for CFAbd-Score between control and CF group

#### PAC-SYM

There was no difference in the total score between the control and CF groups (control median 4 [IQR 1, 7] vs. CF 5 [IQR 2, 7], p=1, Wilcoxon), nor were there any correlations with MRI metrics.

#### Likert scale for flatulence, bloating and abdominal pain

The individual symptom score and total scores using the Likert scale were low for both study groups during the study day. There was no difference in the individual symptoms or total score between the CF and control groups.

### Exploratory outcome

#### Differences in MR images

Qualitative observation of the MR images obtained identified appearances consistent with faecal material (usually seen in the large bowel) within the distal small bowel of CF patients that was not seen in healthy controls or previously seen in similar studies^23,31^ (Supplementary Figure 1).

## Discussion

Our study has shown, for the first time, that MRI methodologies used in other gastrointestinal disorders can elucidate gut function and transit in CF. We have compared people with CF with age and sex-matched controls and demonstrated that the CF group have prolonged OCTT, as well as an increase in both small bowel water and colonic volume compared to controls (corrected for body surface area). For each of these MRI metrics the differences are statistically significant and a prolongation of OCTT of this magnitude is clinically meaningful. The increase in small bowel water which we observed in CF is the opposite of our study hypothesis which predicted that small bowel water would be reduced.

OCTT was delayed in the CF group compared to the control group, in line with our hypothesis. Our data are consistent with a previous report of delayed transit time in CF assessed by wireless motility capsule (SmartPill^®^).^32^ No difference in gastric half-emptying times in CF is also in agreement with the SmartPill^®^ data and suggests that the delayed transit is within the small bowel. Transit time in the control group was similar to a previous study of healthy individuals using MRI.^22^ Our method assesses the normal physiological process by tracking the arrival of food in the caecum. This non-invasive observation contrasts with the SmartPill^®^ which, due to its size, only leaves the stomach when propelled by a migrating motor complex and in association with gastric sieving.^32-34^ The use of the SmartPill^®^ is not recommended in people under 18 years of age.^35^

Whilst delayed small bowel transit time is in agreement with other studies, we cannot exclude an element of delayed gastric emptying. There is limited evidence in previous published studies, which have small sample sizes and use different techniques for measuring gastric emptying times.^36-41^

Delayed transit due to disorders of gastrointestinal motility is associated with nausea, bloating and a predisposition to small intestinal bacterial overgrowth.^42^ The limited evidence base in dysmotility would suggest use of prokinetics but this is not current practice in CF. European guidelines^43^ for distal intestinal obstruction syndrome in CF recommend the osmotic laxative polyethylene glycol (PEG) as first line treatment, although no randomised controlled trials have tested this approach. Treatment with PEG is consistent with the disease paradigm of reduced intestinal water content.

Contrary to our expectations, we observed a higher AUC for corrected SBWC in CF. SBWC was higher throughout the period between meals and notably did not fall immediately after the second challenge meal to the same degree as in the controls. The higher volume of SBWC suggests an altered balance between intestinal secretion, absorption and motility. A previous study between healthy volunteers and people with constipation showed that ingestion of psyllium husk increased small bowel water through its capacity to bind water and prevent absorption. One plausible hypothesis would be that people with CF have an impaired capacity to absorb intestinal water because of their hyper-viscous epithelial mucus.^44^ However, there are two alternative mechanisms, both of which might also explain both increased small bowel water and prolonged OCTT.

The first is suggested by the post-prandial response to the second challenge meal. The gastro-ileal reflex refers to discharge of ileal content into the colon on the arrival of calorific material in the stomach.^45^ In our previous MRI work this has manifested as a substantial fall in SBWC (100-150ml) after a meal.^26^ In our study, we observed a fall in SBWC following the second meal (after scan T240) in the controls. However, this drop was markedly reduced in the CF group. This may reflect restricted flow at the terminal ileum, a recognised site of pathology in CF. Viscous mucus at this pinch point in the bowel may cause partial obstruction such that the chyme held up undergoes excessive desiccation. Such appearances are described in an early surgical case series of distal intestinal obstruction syndrome.^46^ The fact that we have found a transit delay in a sample group where the majority of participants had no history of distal intestinal obstruction syndrome raises the possibility that sub-clinical pathology in the terminal ileum is prevalent in CF patients. Stasis and prolonged transit times in the small bowel may lead to small intestinal bacterial overgrowth and inflammation.^19^

The second possible mechanism is an exaggerated ileal brake, where ileal fat delays jejunal transit.^47^ Inadequate availability of pancreatic enzymes for digestion could lead to increased fat in the terminal ileum and the release of the gut-derived peptide YY (PYY) and glucagon-like peptide-1 (GLP-1) hormone.^47,48^ This would further slow gastric emptying and the expulsion of small bowel contents into the caecum. Investigation of the exact pathology at the terminal ileum further will be an important focus of future research. Either of these mechanisms raises questions about whether an osmotic agent such as PEG is the best approach for CF patients with apparent symptoms of constipation, or whether a more selective approach is needed.

Increases in colonic volumes in the people with CF could reflect the increased prevalence of constipation reported by this group as a similar increase is found in functional constipation with slow transit.^29^ Our raw colonic volume data (Supplementary Table 1, Supplementary Table 2) are consistent with other studies for healthy controls and constipation groups.^23,31,49^

The gastrointestinal tract continues to develop and grow until adulthood.^50^ Although we matched the study groups to age and gender, we expected that there would be a difference in height and weight in view of the nutritional challenges of CF.^51^ Previous studies have described increasing colonic volumes with height^23^ and total intestinal length with height and weight.^52,53^ Correcting data by body surface area, rather than body mass index, is a recognised approach in paediatric care, such as to determine appropriate drug doses^54^, estimate glomerular filtration rates^55^ and cardiac stroke volume.^56^ We therefore opted to adjust for height and weight by dividing colonic volumes and SBWC by body surface area.^50^

We assessed patient symptoms as recent studies have shown a significant difference in gastrointestinal questionnaire scores in a CF group compared to a control group.^14^ This is not the case in our study and may be because we excluded people with previous extensive gut surgery and those unable to stop drugs affecting bowel habit. It may also reflect the small sample size; since studies demonstrating a difference in gastrointestinal symptoms generally require larger sample sizes.^8,11,14,15^ Our study illustrates that objective markers of gut function in CF may be more sensitive in detecting an abnormality than subjective symptom report. Further work is needed to determine whether these methods can also detect response to therapy in a smaller trial than would be needed to confirm effectiveness using a symptom-based outcome.

### Strengths and weaknesses

The advantage of using a completely non-invasive and non-radiation imaging technique is that the gastrointestinal tract is undisturbed to allow a better understanding of the mechanism of disease. Despite concomitant chest disease, people with CF were able to tolerate multiple breath-hold sequences during scanning. This technique is highly appealing to a patient group with high treatment burden.

Our study has been able to capture dynamic changes in the small bowel and to observe transit times through multiple MRI scans. Such a protocol is, however, time consuming and costly, which limits its capacity for dissemination. Our future research using MRI will refine scanning protocols and reduce the number of MRI scans with the aim of implementing this method of assessment in clinical practice.

Another limitation is the need for a trained researcher to evaluate multiple images in order to derive OCTT. In the future, transit time could be more easily measured with new emerging MRI tools such as mini-capsules visible on MRI.^57^ These mini-capsules would be more in keeping with the physiological mechanism of gastric emptying than larger capsules or devices.^58^

### Future directions

Our study has shown abnormalities in MRI metrics of gastrointestinal function and transit in participants with CF compared to matched controls. The direction of future MRI studies should compare CF populations with a history of distal intestinal obstruction syndrome or daily gastrointestinal symptoms with those who report minimal symptoms to further explore the spectrum of gut pathophysiology in CF. The same approach can then be applied to evaluating commonly used therapies designed to prevent distal intestinal obstruction syndrome or relieve symptoms – many of which have little or no clinical trial evidence to support their use in CF, and also new CFTR modulator therapies.

Our gastrointestinal MRI group has observed changes in digestion in a range of patients using an all-day scanning protocol similar to the one described for this study.^26,31,34,59^ We were able to tailor the scanning protocol to accommodate the concomitant lung disease of people with CF. Future clinical trials will be able to use this protocol, and exact time points of scans and meals provided can be tailored to answer specific research questions.

## Conclusions

This is the first study to demonstrate that investigating underlying gut pathologies in CF using MRI is achievable. It has been long known that MRI is safe and non-invasive but introducing a scanning protocol adapted to the breathing requirements of a person with CF is novel.

Our methodology has provided a platform for studying gastrointestinal complications in CF. We have demonstrated that there is a difference in pre-defined outcomes between controls and people with CF which reflect the pathophysiological mechanisms of gut disease. We have identified novel areas of gut function in CF as a target for future research, in particular the reduced SBWC drop after a high calorie meal which could reflect terminal ileum obstruction.

The next steps will require collaboration between the gastroenterology and CF research communities to improve treatment and quality of life for people with CF.

## Data Availability

The original dataset will be made available by the corresponding author on request and after signing a confidentiality agreement not to publicly share the data.

## Abbreviations

(AUC): Area under the curve
(CF): Cystic fibrosis
(CFTR): Cystic fibrosis transmembrane conductance regulator
(GLP-1): Glucagon-like peptide-1
(MRI): Magnetic resonance imaging
(OCTT): Oro-caecal transit time
(PAC-SYM): Patient Assessment of Constipation-Symptoms
(PAGI-SYM): Patient Assessment of Gastrointestinal Disorders Symptom Severity Index
(QoL): Quality of life
(SBWC): Small bowel water content

